# Virtual care use among older immigrant adults in Ontario, Canada during the COVID-19 pandemic: a repeated cross-sectional analysis

**DOI:** 10.1101/2022.07.20.22277848

**Authors:** Janette Brual, Cherry Chu, Jiming Fang, Cathleen Fleury, Vess Stamenova, Onil Bhattacharyya, Mina Tadrous

**Author notes:** **Corresponding author:** Cherry Chu, 76 Grenville Street, Toronto, Ontario M5S 1B2. (<u>co-first author</u>).

## Abstract

The critical role of virtual care during the COVID-19 pandemic has raised concerns about the widening disparities to access by vulnerable populations including older immigrants. This paper aims to describe virtual care use in older immigrant populations residing in Ontario, Canada.

In this population-based, repeated cross-sectional study, we used linked administrative data to describe virtual care and healthcare utilization among immigrants aged 65 years and older before and during the COVID-19 pandemic. Visits were identified weekly from January 2018 to March 2021 among various older adult immigrant populations.

Among older immigrants, over 75% were high users of virtual care (had two or more virtual visits) during the pandemic. Rates of virtual care use increased for both older adult immigrant and non-immigrant populations. At the start of the pandemic, virtual care use was lower among immigrants compared to non-immigrants (weekly average of 77 vs 86 visits). As the pandemic progressed, the rates between these groups became similar (80 vs 79 visits). Virtual care use was consistently lower among immigrants in the family class (75 visits) compared to the economic (82 visits) or refugee (89 visits) classes, and was lower among those who only spoke French (69 visits) or neither French nor English (73 visits) compared to those who were fluent in English (81 visits).

This study found that use of virtual care was comparable between older immigrants and non-immigrants overall, though there may have been barriers to access for older immigrants early on in the pandemic. However, within older immigrant populations, immigration category and language ability were consistent differentiators in the rates of virtual care use throughout the pandemic.

**Author Summary:** When the COVID-19 pandemic began, healthcare systems pivoted from in-person to virtual care to maintain physical distancing. Studies have shown that virtual care use became much more frequent during the pandemic as a result. What we do not know is whether virtual care is being used equitably, that is, whether everybody has fair access to the resource. This can be a big issue particularly amongst older adults, who are often battling several diseases and use healthcare frequently. Many older adults are immigrants who may face challenges in accessing healthcare due to reasons such as limited language fluency and resource support. Our study found that older adult immigrants aged 65 and above living in Ontario, Canada had lower use of virtual care initially, but their use eventually caught up with non-immigrants as the pandemic progressed. We also found that older adult immigrants from the family class had lower virtual care use compared to those from the economic, refugee, or other immigration classes. Additionally, immigrants who were not fluent in English had lower use compared to those who were fluent. These results show that virtual care access remains an issue for vulnerable minorities and steps should be taken to ensure these groups are receiving adequate care.

## Introduction

In March 2020, the World Health Organization declared the outbreak of the novel coronavirus disease (COVID-19) a pandemic[1], and as a result, health systems around the world had to adapt quickly in order to continue providing access to healthcare services. To accommodate safety protocols of physical distancing and to reduce virus transmission, health care shifted from predominantly in-person care to remote care using virtual modalities. In response, the Ontario government issued temporary billing codes allowing physicians to bill for the use of telephone and video visits[2]. A recent study examining the period before and during the COVID-19 pandemic found that virtual care has been maintained at slightly above 50% of all ambulatory visits in Ontario, Canada[3].

As virtual care becomes more common and innovations in digital health continue to be developed post-pandemic, there is growing concern that the digital divide will widen and disadvantage the most vulnerable populations[4, 5], particularly older immigrants. Older adults have complex health needs, higher rates of chronic disease, disabilities, and comorbidities, and are among the highest users of the healthcare system[6, 7]. Older immigrants in Canada are even more vulnerable to breakdowns in the system, and may face additional challenges when accessing healthcare, including cultural differences, discrimination, language barriers, literacy, health beliefs, and spatial isolation[8-11]. In addition, some barriers to access for older adult populations, such as physical or mental disabilities, inexperience or discomfort with technology, or lack of digital equipment[12-14] may be even more pronounced among immigrants who migrated at an older age and have poor social determinants of health upon arriving in Canada. Furthermore, some recent immigrants may face further health decline with increasing years of residency in Canada and poor access to health care[15]. While early reports during the pandemic showed that the rates of virtual care visits were highest among older adults compared to younger groups[16], it remains unclear how immigration status affected virtual care use.

This paper aims to describe virtual care use in older immigrant populations in Ontario overall, as well as across various immigrant sub-groups.

## Methods

### Data Sources

We used linked and coded population-based health databases from ICES (formerly the Institute for Clinical Evaluative Sciences) to examine the utilization of virtual care among the older adult patient population. ICES is an independent, non-profit research institute whose legal status under Ontario’s health information privacy law allows it to collect and analyze health care and demographic data, without consent, for health system evaluation and improvement.

The databases used for our study included: (1) The Immigration, Refugee, and Citizenship Canada (IRCC) Permanent Residents Database, which has demographic data of all immigrants to Canada who become permanent residents including information on country of birth, citizenship and country of residence, admission category and landing date or age at landing; (2) Registered Persons Database (RPDB), which contains demographic information of all patients covered under the Ontario Health Insurance Plan; (3) Ontario Health Insurance Plan (OHIP), which includes information on all health services delivered by physicians to Ontario patients who are eligible for coverage; (4) various ICES-derived cohorts. Databases were linked using unique encoded identifiers and analyzed at ICES.

### Study design and setting

This analysis reports on virtual care use among older immigrant populations and is part of a larger study of virtual care use among older adults before and during the COVID-19 pandemic. The results of the broader study are reported elsewhere[16]. We conducted a population-based, repeated cross-sectional study of virtual ambulatory visits among older immigrants (age 65 years and older) residing in Ontario, Canada with valid Ontario Health Insurance Plan (OHIP) healthcare coverage. Individuals who were non-Ontario residents, had an invalid health card number or were residing in long-term care were excluded from analysis. Visits were identified using OHIP administrative claims data before and during the COVID-19 pandemic, from January 1, 2018 to March 29, 2021. All OHIP billing codes used to identify and calculate the rates of virtual visits are available in the Appendix.

### Patient characteristics

Various subgroups of patients were identified using the relevant databases, including patient demographics and immigration records. ‘Immigrant’ refers to those who immigrated to Canada during or after 1985. ‘Non-immigrant’ subgroups refer to any individual not recorded in the immigration database and includes both Canadian-born residents and immigrants who arrived in Canada before 1985. ‘Recent immigrant’ includes immigrants who arrived during or after 2016 and ‘Earlier immigrant’ includes immigrants who arrived between 1985 and 2015. Among the immigration classes, the family category involves reunification of Canadian citizens or permanent residents with close family members such as spouses, common-law or conjugal partners, and dependent children[17]. Economic immigrants are selected based on the National Occupational Classification according to skills and ability to contribute to the Canadian economy. This category may include immigrants in the business category (entrepreneurs, investors and self-employed), as well as skilled and unskilled workers. The refugee category refers to immigrants who received status for refugee and asylum protection. Finally, the ‘other’ category refers to immigrants and permanent residents who do not belong to the other immigration categories, and may include immigration by humanitarian and compassionate reasons.

We used the Rurality Index of Ontario (RIO)[18] to determine rurality among the study population, comparing urban (RIO score < 40) versus rural (RIO score ≥ 40) residence. The Postal Code Conversion File (PCCF) was used to convert all patient postal codes to neighborhood income quintiles. A number of ambulatory care sensitive conditions (ACSC) were also reported and included chronic obstructive pulmonary disease (COPD), heart failure, asthma, hypertension, angina, and diabetes. Hospitalizations due to ACSC conditions are often used as an indicator of health system performance. ACSC can be managed through timely and effective disease management within primary care[19] and greater comorbidity of these conditions could increase the risk of hospitalization if not well managed.

We further categorized patients based on their use of virtual care (high vs low) to compare health status, immigrant status, and health services utilization. High users were defined as patients who received two or more virtual visits after March 14, 2020 until end of study period, while low users were defined as patients who received one or no virtual visits after March 14, 2020 until end of study period.

### Analysis

Rates of virtual visits were calculated for each week (from Sunday to Saturday) from January 1st, 2018 to March 29th, 2021. The denominator for each week’s rate calculation included all residents of Ontario who were age 65 and above during the week and eligible for healthcare services in Ontario (i.e., OHIP-insured). We summarized the overall rates of virtual visits within the older immigrant population, comparing rates to the non-immigrant population and across the following subgroups: recent versus non-recent immigrant status, immigration category (economic, family, refugee and ‘other’ category), and Canadian language ability (English and French). For this analysis, March 14, 2020 was considered the start date of the COVID-19 pandemic as it was the day that new temporary billing codes were introduced by the Ontario government which expanded physician reimbursement of telemedicine services (including telephone calls) in response to the pandemic. All analyses were performed in SAS 9.4 (SAS Institute)[20].

### Ethics Approval

We received approval from the Women’s College Hospital Research Ethics Board for use of the IRCC – Permanent Residents Database. Ethics review through a research ethics board was not required for use of the other administrative databases for the purposes of this study as authorized under section 45 of Ontario’s Personal Health Information Protection Act[21].

## Results

### Overall patient characteristics by immigration status

Baseline patient characteristics are summarized in **Table 1** comparing older adult immigrants and non-immigrants. Overall, a greater proportion of older adults were women compared to men (54.2% versus 45.8%). Immigrants were younger with 73.7% compared to 66.6% of non-immigrants in the youngest-old category (65-74 years). A higher proportion of older immigrants lived in urban areas compared to non-immigrants (99.0% versus 89.1%), and they had lower incomes, with 26.3% of older immigrants compared to 18.0% of non-immigrant older adults in the lowest income quintile. For both immigrant and non-immigrant patient populations, the top previously diagnosed disease conditions were hypertension, diabetes, and mental health. The number of patients with hypertension and mental health was similar between these two groups, however, more immigrants had diabetes compared to non-immigrants (42% versus 30%).

**Table 1.** Patient characteristics for older immigrant and non-immigrant adults in Ontario, Canada.

| Characteristic | Total<br>N=2,282,798 | Immigrant<br>n=271,075 | Non-Immigrant<br>n=2,011,723 | Standardized<br>Difference<sup>a</sup> |
| --- | --- | --- | --- | --- |
| Age, n(%) |  |  |  |  |
| 65-74 | 1,539,340 (67.4) | 199,780 (73.7) | 1,339,560 (66.6) | 0.156 |
| 75-84 | 567,625 (24.9) | 57,168 (21.1) | 510,457 (25.4) | 0.102 |
| 85+ | 175,833 (7.7) | 14,127 (5.2) | 161,706 (8.0) | 0.114 |
| Sex, n(%) |  |  |  |  |
| Female | 1,233,462 (54.0) | 146,974 (54.2) | 1,086,488 (54.0) | 0.004 |
| Male | 1,049,336 (46.0) | 124,101 (45.8) | 925,235 (46.0) | 0.004 |
| Neighbourhood<br>income quintile,<br>n(%) |  |  |  |  |
| 1 | 432,945 (19.0) | 71,331 (26.3) | 361,614 (18.0) | 0.202 |
| 2 | 469,631 (20.6) | 61,869 (22.8) | 407,762 (20.3) | 0.062 |
| 3 | 458,824 (20.1) | 56,250 (20.8) | 402,574 (20.0) | 0.018 |
| 4 | 439,791 (19.3) | 46,581 (17.2) | 393,210 (19.5) | 0.061 |
| 5 | 476,247 (20.9) | 34,560 (12.7) | 441,687 (22.0) | 0.245 |
| Rurality, n(%) |  |  |  |  |
| Urban | 2,060,282 (90.3) | 268,240 (99.0) | 1,792,042 (89.1) | 0.426 |
| Rural | 200,182 (8.8) | 2,203 (0.8) | 197,979 (9.8) | 0.41 |
| Region, n(%) |  |  |  |  |
| Central | 690,994 (30.3) | 154,034 (56.8) | 536,960 (26.7) | 0.642 |
| East | 599,098 (26.2) | 56,981 (21.0) | 542,117 (26.9) | 0.139 |
| North | 144,377 (6.3) | 1,119 (0.4) | 143,258 (7.1) | 0.358 |
| Toronto | 174,752 (7.7) | 26,179 (9.7) | 148,573 (7.4) | 0.081 |
| West | 673,577 (29.5) | 32,762 (12.1) | 640,815 (31.9) | 0.492 |
| Chronic<br>conditions, n(%) |  |  |  |  |
| Hypertension | 1,583,965 (69.4) | 194,053 (71.6) | 1,389,912 (69.1) | 0.055 |
| Diabetes | 721,699 (31.6) | 114,210 (42.1) | 607,489 (30.2) | 0.25 |
| Mental Health | 751,608 (32.9) | 79,545 (29.3) | 672,063 (33.4) | 0.088 |
| Asthma | 223,451 (9.8) | 23,670 (8.7) | 199,781 (9.9) | 0.041 |
| Angina | 166,430 (7.3) | 20,158 (7.4) | 146,272 (7.3) | 0.006 |
| CHF | 202,396 (8.9) | 17,881 (6.6) | 184,515 (9.2) | 0.096 |
| COPD | 188,974 (8.3) | 11,536 (4.3) | 177,438 (8.8) | 0.185 |
| Dementia | 68,528 (3.0) | 6,934 (2.6) | 61,594 (3.1) | 0.03 |

<sup>a</sup> Standardized difference greater than 0.1 is considered statistically significant.

### Immigrant characteristics by high vs low virtual care user group

Among immigrants residing in Ontario, most arrived under the family immigration category (50.4%), followed by immigrants under the economic category (32.7%) with the fewest in the refugee and ‘other’ immigration categories (13.7% and 3.1%, respectively) (**Table 2**). In terms of Canadian language ability (English and French), more than half of immigrants reported being able to speak English (53.3%), followed by immigrants who spoke neither English nor French (44.3%). Few immigrants were able to speak both English and French (1.6%) or French only (0.8%). Most immigrants arrived before January 1^st^, 2016 (97.8%) with few recent immigrants (2.2%). 78% of the immigrant population were high users of virtual care during the COVID-19 pandemic. The mean number of ACSC was 1.50±0.98 and 1.09±0.91 in high and low users of virtual care, respectively. The average number of outpatient visits was higher among high users (3.81±4.37) compared to low users (1.42±3.18). Similarly, more high virtual care users had at least one ED visit (13.5% vs 8.2%) and hospitalization (4.8% vs 2.8%) than low users.

**Table 2:** Patient characteristics and healthcare utilization among older immigrant adults only, by virtual care use in Ontario, Canada.

| Virtual Care Group<sup>a</sup> | Total<br>N=271,075 | High User<br>N=211,304 | Low User<br>N=59,771 | Standardized<br>Difference<sup>c</sup> |
| --- | --- | --- | --- | --- |
| Immigration Category, n (%) |  |  |  |  |
| Family | 136,755 (50.4) | 108,926 (51.5) | 27,829 (46.6) | 0.1 |
| Economic | 88,608 (32.7) | 67,672 (32.0) | 20,936 (35.0) | 0.064 |
| Refugee | 37,144 (13.7) | 28,109 (13.3) | 9,035 (15.1) | 0.052 |
| Other | 8,568 (3.2) | 6,597 (3.1) | 1,971 (3.3) | 0.01 |
| Immigrant Language Ability, n (%) |  |  |  |  |
| Both French and English | 4,377 (1.6) | 3,375 (1.6) | 1,002 (1.7) | 0.006 |
| English | 144,479 (53.3) | 113,001 (53.5) | 31,478 (52.7) | 0.016 |
| French | 2,034 (0.8) | 1,480 (0.7) | 554 (0.9) | 0.025 |
| Neither French nor English | 120,019 (44.3) | 93,304 (44.2) | 26,715 (44.7) | 0.011 |
| Not Stated | 166 (0.1) | 144 (0.1) | 22 (0.0) | 0.014 |
| Time of Arrival in Canada<sup>b</sup>, n (%) |  |  |  |  |
| Earlier immigrant | 265,233 (97.8) | 206,674 (97.8) | 58,559 (98.0) | 0.011 |
| Recent immigrant | 5,842 (2.2) | 4,630 (2.2) | 1,212 (2.0) | 0.011 |
| Number of ACSC, mean (SD) | 1.41 (0.98) | 1.50 (0.98) | 1.09 (0.91) | 0.425 |
| Number of virtual care visits, mean (SD) | 5.94 (6.30) | 7.45 (6.37) | 0.6 (0.49) | 1.518 |
| Number of outpatient visits in 6 months prior, mean (SD) | 3.28 (4.25) | 3.81 (4.37) | 1.42 (3.18) | 0.625 |
| Visited ED in 6 months prior, n(%) | 33,456 (12.3%) | 28,561 (13.5%) | 4,895 (8.2%) | 0.172 |
| Hospitalized in 6 months prior, n(%) | 11,831 (4.4%) | 10,180 (4.8%) | 1,651 (2.8%) | 0.108 |

<sup>a</sup> High virtual care use is defined as two or more virtual visits after March 14, 2020, and low users, defined as zero or one virtual visits after March 14, 2020.
<sup>b</sup> "Recent immigrant" refers to immigrants who arrived in Canada after Jan 1, 2016.
<sup>c</sup> Standardized difference greater than 0.1 is considered statistically significant.

**Fig 1** compares weekly rates of virtual care use between older immigrant and non-immigrant populations. Both reported a significant increase in the rates of virtual care visits at the start of the COVID-19 pandemic, but the rates in non-immigrants were greater than those for immigrants, with average weekly rates (per 1000) of 86 and 77, respectively. However, as the pandemic continued onward, virtual care use between both groups became similar, with average rates (per 1000) of 79 and 80, respectively.

**Fig 1.**
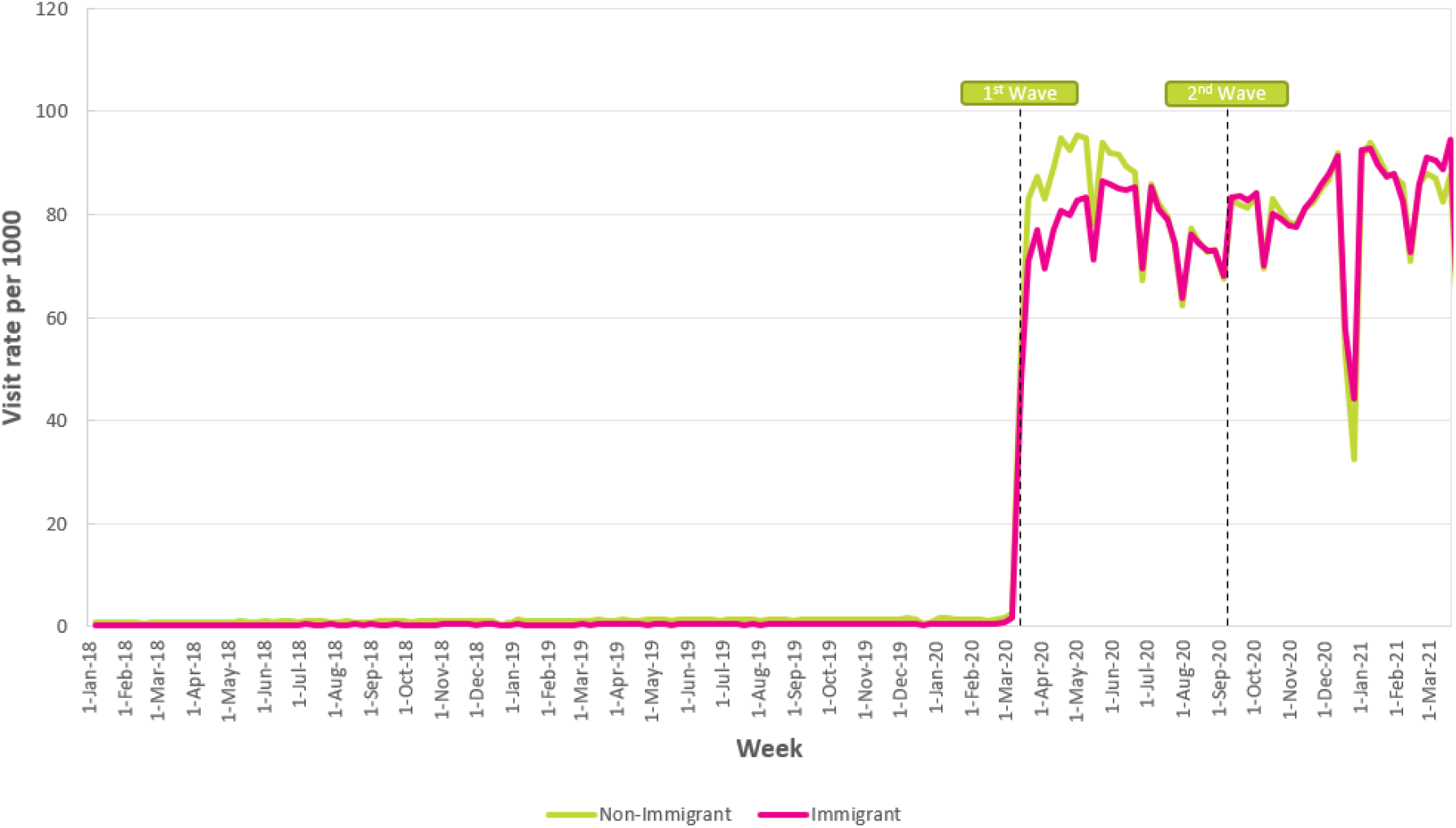
Rate of virtual visits per 1000 eligible older adult patients in Ontario: Immigrant versus non-immigrant subgroups, 2018-2021. *Note: Non-immigrants may include immigrants who arrived in Canada before 1985*

### Virtual Care Use by Immigration Status

As shown in **Fig 2**, during the first wave of the pandemic, earlier immigrants had slightly higher rates of virtual care use compared to recent immigrants (arrived after January 1, 2016), with average weekly rates (per 1000) of 78 and 71, respectively, although both groups had lower rates compared to non-immigrants (86 visits per 1000). By the summer of 2020 and onwards into the pandemic, rates for both earlier and recent immigrants increased to similar rates as non-immigrants, with average weekly rates (per 1000) of 80, 78, and 79, respectively.

**Fig 2.**
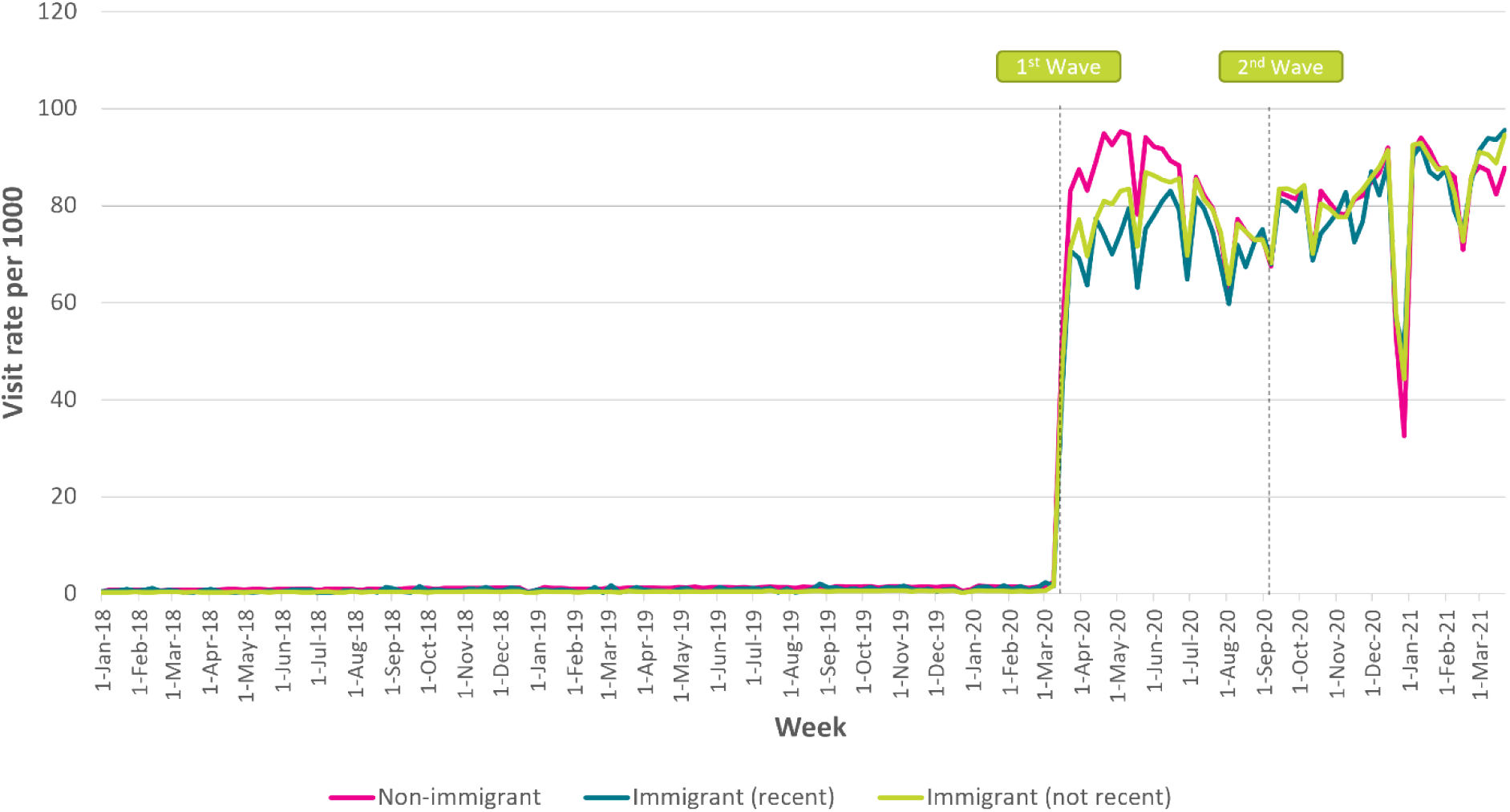
Rate of virtual visits per 1000 eligible older immigrant patients in Ontario by recency of immigration, 2018-2021. *Note: Recent=immigrated after Jan 1, 2016; Non-immigrants may include immigrants who arrived in Canada before 1985*

### Virtual Care Use by Canadian Language Ability

**Fig 3** shows weekly rates of virtual care use according to Canadian language ability among non-immigrant and immigrant subgroups. All groups reported a significant increase in weekly rates of virtual care visits at the start of the COVID-19 pandemic. From the first wave onwards into the pandemic, virtual care use was highest among non-immigrants, immigrants with English language ability, and immigrants who speak both French and English, with average weekly rates (per 1000) of 81, 86, and 79, respectively. However, those who speak neither English nor French or only speak French had lower virtual care use, with average weekly rates (per 1000) of 69 and 73, respectively.

**Fig 3.**
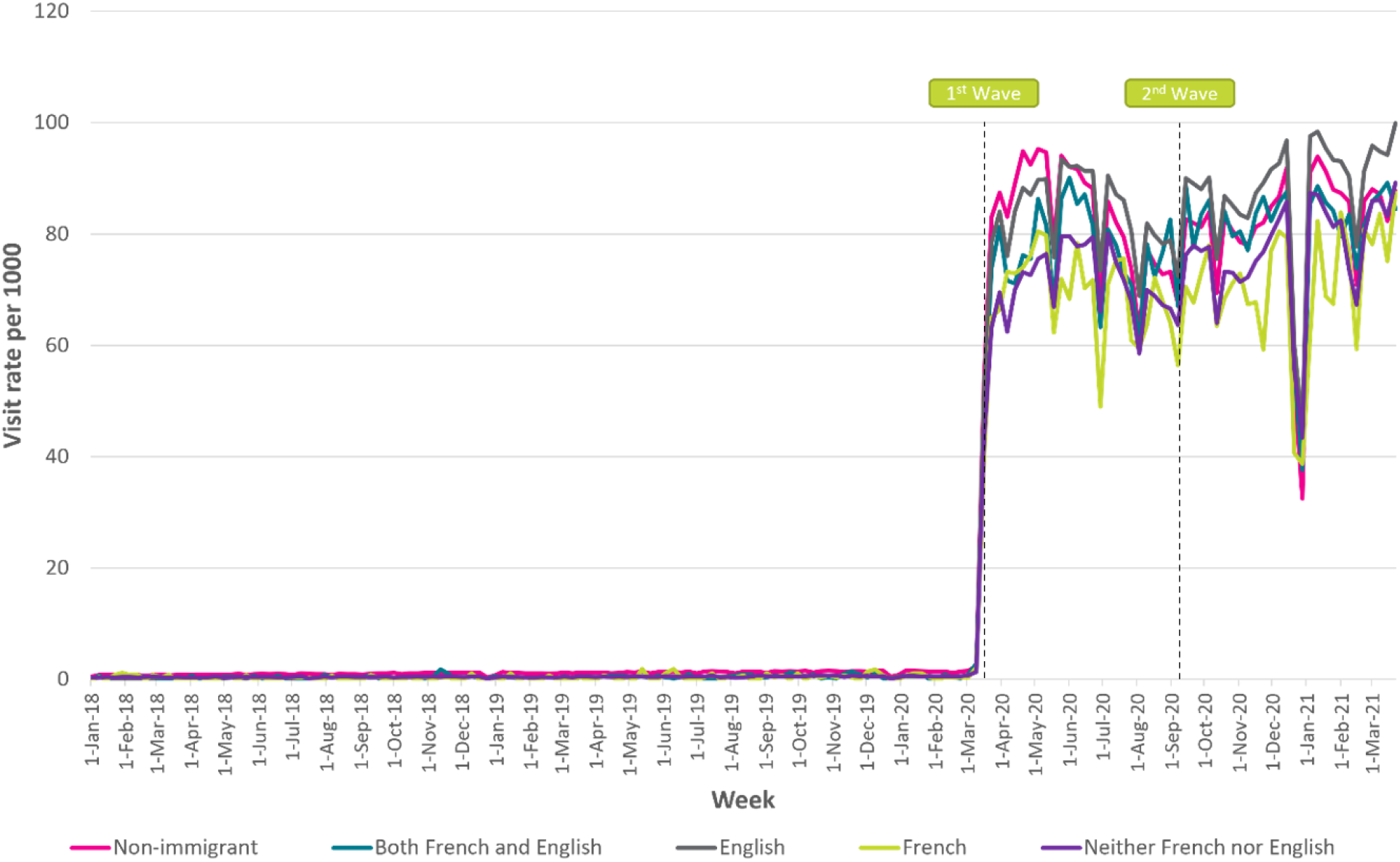
Rate of virtual visits per 1000 eligible older immigrant patients in Ontario by Canadian language ability, 2018-2021. *Note: Non-immigrants may include immigrants who arrived in Canada before 1985*

### Virtual Care Use by Immigration Category

All immigration categories reported a significant increase in weekly rates of virtual care visits during the COVID-19 pandemic as shown in **Fig 4**. From the first wave onwards, virtual care use was highest among refugees and those in the ‘other’ immigration categories, with average weekly rates (per 1000) of 89 for both. These rates were followed by non-immigrants and immigrants in the economic immigration category (e.g., skilled and unskilled workers), with average weekly rates (per 1000) of 81 and 82, respectively. Older immigrants in the family category used virtual care the least, with average weekly rates (per 1000) of 75.

**Fig 4.**
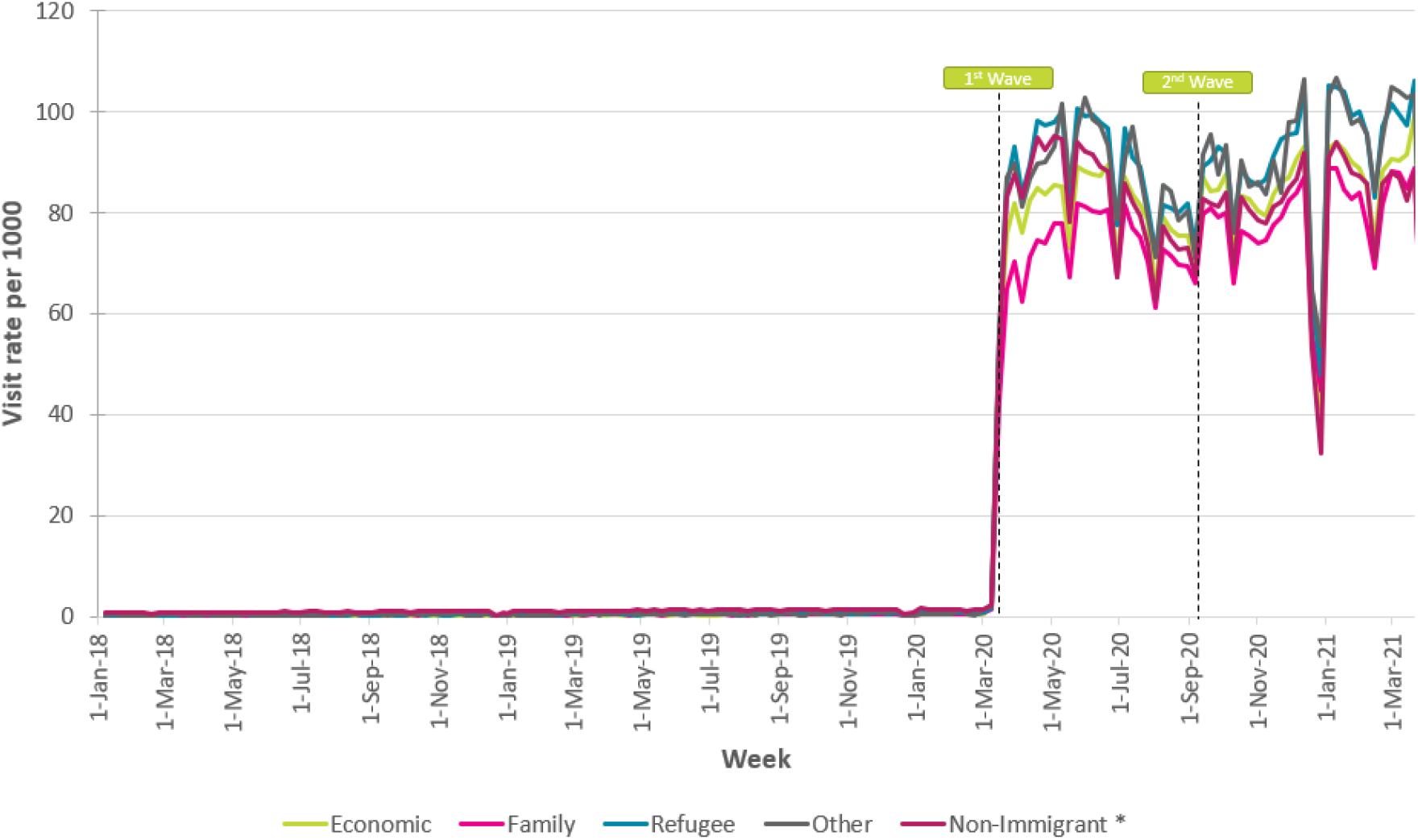
Rate of virtual visits per 1000 eligible older immigrant patients in Ontario by immigration category, 2018-2021. *Note: Non-immigrants may include immigrants who arrived in Canada before 1985*

## Discussion

Overall, this study showed that virtual care use was lower among older immigrant populations when the pandemic began compared to non-immigrant populations - however, as the pandemic progressed the rates between these groups converged and became similar. Among older immigrant populations, immigration admission category and language ability were found to be consistent differentiators in the rates of virtual care use throughout the pandemic in Ontario. Despite the consistent finding of increased virtual care uptake across all demographic groups assessed, there remains potential equity issues with adoption within the older adult immigrant population.

While there was an overall reduction in health service use at the start of the pandemic as health systems shifted to virtual methods, there was a lag in virtual care adoption among older immigrant populations. Of particular concern are older immigrants who had more chronic or severe health issues, where having limited access to care at the start of the pandemic may have had detrimental effects on their health resulting in greater use of care as the pandemic progressed. Reasons for this apparent lagged difference in virtual care uptake for immigrants is unknown, however these findings suggest that although there are increasing healthcare needs among older immigrant populations, there may be significant barriers when accessing virtual care services. Some of these barriers may be related to language, socioeconomic status, and other immigrant-related experiences.[8-11]

Within the immigrant cohort, virtual care use was lowest among those in the family immigration category when compared to other categories, while virtual care use was higher among the refugee and economic classes of older immigrants. Lower rates of virtual care use among older immigrants of the family class was a surprising finding, as individuals in this group, by definition, would likely have family members or strong social support to assist them with attending virtual visits. However, shifts in family dynamics over time (death of spouse, children moving with the labour market, transitional care from home-setting, etc.), particularly among earlier immigrants, can impact the availability of economic and social resources in later life, resulting in poorer access to care[22]. Another possible explanation is that many newer immigrant families may be of lower socioeconomic status[23], which could present as a barrier to accessing virtual care, particularly with limited access to internet or digital tools (i.e. digital device), in addition to non-technical barriers such as digital health literacy, comfort with digital technology, and lack of trust towards digital devices[24]. Economic class immigrants reported the second lowest rates of virtual care use – although the reason for this finding is unclear, it may be related to these individuals being younger and healthier than other immigrants. Canadian immigration and settlement are not monolithic experiences, and the intersectionality of realities experienced by immigrants during the pandemic and onward can greatly impact their access to health care services overall, including virtual care.

Our analyses also found that the lowest rates of virtual care visits during the pandemic were among immigrants who only spoke French, and immigrants who had neither English nor French-language proficiencies. Immigrants who are unable to communicate in the dominant language can experience several issues relating to patient safety, appropriate treatment, and quality of care[25]. The provision of healthcare services in French (outside of Quebec) and non-official or minority languages is a significant resource challenge, and many immigrants experience difficulties understanding their healthcare providers, and may find the quality of translation services inconsistent or availability of services unreliable[26]. Ad hoc interpreters and use of digital translation tools such as Google translate, while helpful, may not be adequate for patients to receive the same quality of care. A few studies have cited the impact of language barriers in impeding the uptake of virtual care.[27-29] Many of the communication challenges faced by patients who are unable to speak the dominant language in healthcare during in-person encounters likely translate to or worsen with virtual care, regardless of modality, i.e. telephone or video. However, if used optimally, virtual care could alternatively provide a means to connect patients with a healthcare provider who are proficient in their language. Findings from this study show that virtual care provided comparable access to healthcare for many immigrant groups and should be continued as an option for care, but efforts should be made to leverage virtual modalities to decrease the equity gap in access rather than widen it.

## Limitations

There are a few limitations that should be addressed. First, the lack of clinical information that accompanies the use of administrative data requires acknowledgment. Lack of patient-level medical history or previous healthcare use limits our ability to contextualize patients’ use of virtual care as it relates to their health care needs. Second, with the introduction of new COVID-19 virtual care billing codes in Ontario, reimbursement was permitted for both telephone and video visits but did not allow for distinguishing modality used during the visits. As such, we were unable to determine whether the virtual visits identified from the database were delivered via telephone or video. Third, our single factor analyses were unable to examine the intersectionality of the immigrant experience and its association with virtual care access. Therefore, while there may be significant barriers for individuals who are a recent immigrant, non-English speaker, and have low socioeconomic status, we were unable to show how these intersecting identities may impact their use of virtual care. Fourth, we linked to the IRCC Permanent Residents database to identify immigrant status, however this database only includes immigrant data from 1985 onward. Any persons who immigrated to Ontario prior to 1985 would be excluded from the database and as such, we acknowledge that the use of the term “non-immigrant” may also include people who arrived in Ontario before 1985[30]. Similarly, our definition of “recent immigrant” as arriving after Jan 1, 2016 at the time of analysis, may not be an accurate representation of landing experiences in Canada, particularly for immigrants who migrated at an older age. However, the literature supports the notion that immigrants living in Canada for at least 15 to 20 years have similar health statuses and behaviours as Canadian-born residents.[31, 32] Fifth, we were unable to account for inter-provincial migration of immigrants, as the IRCC Permanent Residents database excludes records of immigrants who first landed in a province other than Ontario. Inter-provincial migration can be influenced by different regional economic, political and social factors, as well as being a function of age, behavioural, and lifestyle preferences.[33] However, older immigrants – particularly family class immigrants – are least likely to migrate after arrival in Canada[34], and we can assume inter-provincial migration would be relatively low among this cohort, since selective internal migration often occurs among immigrants of working age. Lastly, the analyses here looked at the short-term shift in virtual care use among older immigrant populations as a result of the COVID-19 pandemic, however future evaluations should continue to monitor virtual care use in older immigrants post-pandemic.

## Conclusion

To our knowledge, this is the first study that examines virtual care use among older immigrant populations, particularly during the COVID-19 pandemic. Our results suggest that there is a clear link between immigration status and virtual care use. In response to the pandemic, public health measures facilitated the rapid uptake of virtual care among the total older adult population, and while there was a lagged difference in use between immigrant and non-immigrant populations in the early months of the pandemic, these rates converged as time progressed. These results suggest that virtual care was able to address gaps in access for older adults broadly, but that the adjustment period from in-person to virtual care, and the learning curve among older immigrant groups, may be longer. Future research should focus on the early access barriers to virtual care that are experienced by vulnerable subgroups such as recent immigrants, as this has important policy implications in a country with many immigrants such as Canada. Additionally, the impact of language barriers and opportunities on the uptake of virtual care should be further researched.

## Data Availability

The data sets generated during and/or analyzed during the current study are not publicly available due to restricted data sharing agreements with ICES and CIHI. Access to the data may be granted by contacting ICES.

## Acknowledgements

This study was supported by ICES, which is funded by an annual grant from the Ontario Ministry of Health (MOH) and the Ministry of Long-Term Care (MLTC). Parts of this material are based on data and information compiled and provided by MOH and the Canadian Institute for Health Information (CIHI). Parts or whole of this material are based on data and/or information compiled and provided by Immigration, Refugees and Citizenship Canada (IRCC). The analyses, conclusions, opinions and statements expressed herein are solely those of the authors and do not reflect those of the funding or data sources; no endorsement is intended or should be inferred.

## References

1. World Health O. WHO Director-General’s Opening Remarks at the Media Briefing on Covid-19 - 11 March 2020. 2020.

2. Ontario Ministry of Health and Long-Term Care. OHIP – Bulletins – Health Care Professionals – MOHLTC.

3. Stamenova V, Chu C, Pang A, Fang J, Shakeri A, Cram P, et al. Virtual care use during the COVID-19 pandemic and its impact on healthcare utilization in patients with chronic disease: A population-based repeated cross-sectional study. PloS one. 2022;17(4):e0267218.

4. Ramsetty A, Adams C. Impact of the digital divide in the age of COVID-19. Journal of the American Medical Informatics Association. 2020;27(7):1147–8.

5. Gray DM, Joseph JJ, Olayiwola JN. Strategies for digital care of vulnerable patients in a COVID-19 world—keeping in touch. 2020;1:e200734–e.

6. Smith B, Magnani JW. New technologies, new disparities: The intersection of electronic health and digital health literacy. Int J Cardiol. 2019;292:280-2. Epub 2019/06/07. doi: 10.1016/j.ijcard.2019.05.066. PubMed PMID: 31171391; PubMed Central PMCID: PMCPMC6660987.

7. Kim EH, Stolyar A, Lober WB, Herbaugh AL, Shinstrom SE, Zierler BK, et al. Challenges to using an electronic personal health record by a low-income elderly population. J Med Internet Res. 2009;11(4):e44. Epub 2009/10/29. doi: 10.2196/jmir.1256. PubMed PMID: 19861298; PubMed Central PMCID: PMCPMC2802566.

8. Wang L, Guruge S, Montana G. Older Immigrants’ Access to Primary Health Care in Canada: A Scoping Review. Canadian Journal on Aging/La Revue Canadienne Du Vieillissement. 2019;38(2):193–209.

9. Christensen LF, Moller AM, Hansen JP, Nielsen CT, Gildberg FA. Patients’ and providers’ experiences with video consultations used in the treatment of older patients with unipolar depression: A systematic review. Journal of Psychiatric and Mental Health Nursing. 2020;27(3):258-71. doi: https://doi.org/10.1111/jpm.12574.

10. Ware P, Bartlett SJ, Paré G, Symeonidis I, Tannenbaum C, Bartlett G, et al. Using eHealth Technologies: Interests, Preferences, and Concerns of Older Adults. Interact J Med Res. 2017;6(1):e3. Epub 2017/03/25. doi: 10.2196/ijmr.4447. PubMed PMID: 28336506; PubMed Central PMCID: PMCPMC5383803.

11. Kruse C, Fohn J, Wilson N, Nunez Patlan E, Zipp S, Mileski M. Utilization Barriers and Medical Outcomes Commensurate With the Use of Telehealth Among Older Adults: Systematic Review. JMIR Med Inform. 2020;8(8):e20359. Epub 2020/08/14. doi: 10.2196/20359. PubMed PMID: 32784177; PubMed Central PMCID: PMCPMC7450384.

12. Hyman A, Stacy E, Mohsin H, Atkinson K, Stewart K, Lauscher HN, et al. Barriers and Facilitators to Accessing Digital Health Tools Faced by South Asian Canadians in Surrey, British Columbia: Community-Based Participatory Action Exploration Using Photovoice. Journal of Medical Internet Research. 2022;24(1):e25863.

13. Jang Y, Chiriboga DA, Molinari V, Roh S, Park Y, Kwon S, et al. Telecounseling for the Linguistically Isolated: A Pilot Study with Older Korean Immigrants. The Gerontologist. 2014;54(2):290–6.

14. Pham Q, El-Dassouki N, Lohani R, Jebanesan A, Young K. The Future of Virtual Care for Older Ethnic Adults Beyond the COVID-19 Pandemic. Journal of medical Internet research. 2022;24(1):e29876.

15. Subedi RP, Rosenberg MW. Determinants of the Variations in Self-Reported Health Status Among Recent and More Established Immigrants in Canada. Social Science & Medicine. 2014;115:103–10.

16. Chu C, Brual J, Fang J, Fleury C, Stamenova V, Bhattacharyya O, et al. The Use of Telemedicine in Older-Adults During the COVID-19 Pandemic: A Weekly Cross-sectional Analysis in Ontario, Canada. Canadian Geriatrics Journal. 2022.

17. Immigration Refugees and Citizenship Canada. Non-Economic Classes. 2018.

18. Kralj B. Measuring Rurality - Rio2008 Basic: Methodology and Results. 2009.

19. Wallar LE, Rosella LC. Risk Factors for Avoidable Hospitalizations in Canada Using National Linked Data: A Retrospective Cohort Study. Plos One. 2020;15(3):e0229465.

20. SAS Statistical Software. 9.4 ed. Cary, NC.: SAS Institute Inc.; 2013.

21. Government of Ontario. Personal Health Information Protection Act, 2004, SO 2004, c. 3, Sched. A. 2014.

22. Leigh JP. Skilled immigrants and the negotiation of family relations during settlement in Calgary, Alberta. Journal of International Migration and Integration. 2016;17(4):1065–83.

23. Ng E, Wilkins R, Gendron F, Berthelot J. Healthy today, healthy tomorrow? Findings from the national population health survey. Dynamics of Immigrants’ Health in Canada: Evidence from the National Population Health Survey, Statistics Canada. 2005.

24. Scott Kruse C, Karem P, Shifflett K, Vegi L, Ravi K, Brooks M. Evaluating barriers to adopting telemedicine worldwide: a systematic review. Journal of telemedicine and telecare. 2018;24(1):4–12.

25. Bowen S. The Impact of Language Barriers on Patient Safety and Quality of Care. Société Santé En Français. 2015:603–23.

26. Ariste R, di Matteo L. Non-Official Language Concordance in Urban Canadian Medical Practice: Implications for Care during the COVID-19 Pandemic. Healthcare Policy= Politiques de Sante. 2021;16(4):84–96.

27. Liang S-Y, Richardson M, Chen T, Colocci N, Kurian A, de Briun M, et al. Widening cancer care disparities in the adoption of telemedicine during COVID 19: who is left behind? Gynecologic Oncology. 2021;162:S23. doi: https://doi.org/10.1016/S0090-8258(21)00690-9.

28. Abdel-Rahman O. Patient-related barriers to some virtual healthcare services among cancer patients in the USA: a population-based study. Journal of Comparative Effectiveness Research. 2021;10(2):119-26. doi: 10.2217/cer-2020-0187.

29. Blackstone SR, Hauck FR. Telemedicine Use in Refugee Primary Care: Implications for Care Beyond the COVID-19 Pandemic. Journal of immigrant and minority health. 2022. doi: 10.1007/s10903-022-01360-6.

30. Chiu M, Lebenbaum M, Lam K, Chong N, Azimaee M, Iron K, et al. Describing the linkages of the immigration, refugees and citizenship Canada permanent resident data and vital statistics death registry to Ontario’s administrative health database. BMC Med Inform Decis Mak. 2016;16(1):135-. doi: 10.1186/s12911-016-0375-3. PubMed PMID: 27769227.

31. Chiu M, Austin PC, Manuel DG, Tu JV. Cardiovascular Risk Factor Profiles of Recent Immigrants Vs Long-Term Residents of Ontario: A Multi-Ethnic Study. Canadian Journal of Cardiology. 2012;28(1):20–6.

32. Perez CE. Health Status and Health Behaviour Among Immigrants [canadian Community Health Survey-2002 Annual Report]. Health Reports. 2002;13:89.

33. Moore EG, Rosenberg MW. Modelling migration flows of immigrant groups in Canada. Environment and planning A. 1995;27(5):699–714.

34. Newbold B. Secondary migration of immigrants to Canada: an analysis of LSIC wave 1 data. The Canadian Geographer/Le Géographe canadien. 2007;51(1):58–71.

